# Racial Differences in Associations Between Adverse Childhood Experiences and Physical, Mental, and Behavioral Health

**DOI:** 10.1101/2023.06.02.23290905

**Authors:** Tracy Lam-Hine, Corinne A. Riddell, Patrick T. Bradshaw, Michael Omi, Amani M. Allen

**Author notes:** **Corresponding Author:** Tracy Lam-Hine, |, Stanford University School of Medicine, Department of Epidemiology & Population Health, 1701 Page Mill Road, Palo Alto, CA 94304.

## Abstract

**Introduction:** Adverse childhood experiences (ACEs) are associated with poor adulthood health, with individuals experiencing multiple ACEs at greatest risk. Multiracial people have high mean ACEs scores and elevated risk of several outcomes, but are infrequently the focus of health equity research. This study aimed to determine whether this group should be targeted for prevention efforts.

**Methods:** We analyzed Waves 1 (1994-95), 3 (2001-02), and 4 (2008-09) of the National Longitudinal Study of Adolescent to Adult Health (n = 12,372) in 2023, estimating associations between four or more ACEs and physical (metabolic syndrome, hypertension, asthma), mental (anxiety, depression), and behavioral (suicidal ideation, drug use) outcomes. We estimated risk ratios for each outcome in modified Poisson models with a race × ACEs interaction, adjusted for hypothesized confounders of the ACE-outcome relationships. We used the interaction contrast to estimate excess cases per 1,000 individuals for each group relative to Multiracial participants.

**Results:** Excess case estimates of asthma were significantly smaller for White (−123 cases, 95% CI: -251, -4), Black (−141, 95% CI: -285, -6), and Asian (−169, 95% CI: -334, -7) participants compared to Multiracial participants. Black (−100, 95% CI: -189, -10), Asian (−163, 95% CI: -247, -79) and Indigenous (−144, 95% CI: -252, -42) participants had significantly fewer excess cases of and weaker (p < 0.001) relative scale association with anxiety compared to Multiracial participants.

**Conclusions:** Adjusted associations between ACEs and asthma or anxiety appear stronger for Multiracial people than other groups. ACEs are universally harmful but may contribute disproportionately to morbidity in this population.

## Background

Adverse childhood experiences (ACEs), also referred to as childhood or early life adversity, are traumatic events during childhood and adolescence that are linked to poor health in adulthood, including five (heart disease, cancer, respiratory diseases, diabetes, and suicide) of the ten leading causes of death in the United States.^1–3^ Over 60% of the US population reports at least one ACE, with up to a third reporting household emotional abuse, parental divorce, or substance abuse, specifically.^4,5^ While children exposed to ACEs may show immediate signs of distress, the health impacts of ACEs may manifest over time, as exposure to trauma early in life predisposes individuals to stressful situations later in life through what has been termed the stress proliferation chain.^6–9^ Trauma is theorized to embed biologically along at least three distinct pathways to influence health: (1) triggering of the biological stress response and resulting impacts on the neuroimmune-endocrine axis,^10,11^ (2) epigenetic expression,^12–14^ and (3) changes in health behaviors due to adaptation or coping with severe or long-term stress.^15–18^

Racial disparities in ACE scores are well-documented, with Multiracial and American Indian/Native American (AI/NA) populations reporting the highest mean ACE score of any racial group.^4,19–22^ In addition to higher ACE scores, Multiracial people report high prevalence of health conditions traditionally linked with ACEs, and poor health in several other conditions. Some studies suggest Multiracial adolescents have poor sleep, behavioral and mental health.^23–29^ Multiracial adults experience the highest prevalence of heart diseases, asthma, obesity, hopeless feelings, and serious psychological distress of any racial group^30–33^, and are significantly more likely to be living in poverty, uninsured, and in poor physical health than monoracial White people.^34,35^ Despite these patterns and the growing size of the Multiracial population, investigators frequently continue to reclassify Multiracial participants into a catchall “other” racial category, existing monoracial categories, or exclude them completely, effectively masking disparities affecting this group.^36,37^

To our knowledge, only two ACEs studies^38,39^ have estimated associations specifically for Multiracial populations. Hall et al’s^38^ study of asthma found that Multiracial participants had the highest mean ACE score and asthma prevalence of any racial group, but no differences in race-specific interaction odds ratios from the reference group (Asians). LaBrenz et al^39^ modeled days of poor mental and physical health, and estimated additive interactions between race and ACE scores. Interaction terms in the mental health model were positive for the Black, Asian, Pacific Islander, Multiracial and “Other” racial groups compared to Whites. For poor physical health days, interaction effect estimates were close to zero and less precise. One small study of monoracial White and Black adolescents in the US South found a stronger dose-response relationship between ACE score and depressive symptoms scores for Black compared to White participants.^40^ Other studies examining monoracial adolescents^41^ and adults^42,43^ have generally found no moderation of health outcomes by race. These findings suggest that the strength of some ACE-health outcome relationships may vary across racial groups, with the direction depending on the referent racial group.

We leverage a nationally representative sample of young adults to determine if the association between ACEs and a range of physical, behavioral, and mental health outcomes are stronger for Multiracial people, and by extension, if prevention efforts should target this group.

## Methods

### Study Sample

The National Longitudinal Study of Adolescent to Adult Health (Add Health) is a longitudinal, nationally representative study following over 20,000 individuals enrolled in grades 7-12 in 1994-95 through four waves of follow-up (1996, 2001-02, 2008-09, 2016-18).^44^ Eighty middle and high schools were selected non-randomly for size, type, grade range, setting, demographics, and geographic location. Students were sampled randomly without stratification from these schools’ enrollment rosters and invited to complete an at-home interview during Wave 1 (n=20,745) which asked questions about the adolescent’s demographics, family background, social networks, home and school environments, and health behaviors. Wave 3 (n=15,197) collected additional life experience and medication information. Wave 4 (n=15,701) was conducted when participants were in their late twenties and included measurements of participants’ metabolic and cardiovascular function.

We categorized participants into five racial groups: White alone, Black alone, Asian alone, American Indian/Native American (AI/NA) alone, (hereafter White, Black, Asian, or AI/NA) or Multiracial. Waves 1 and 3 asked participants to self-identify their race; we used Wave 3 race unless a participant identified as Multiracial at Wave 1 but not Wave 3, in which case we classified them as Multiracial. We excluded Hispanic/Latino participants because race and Hispanic ethnicity are assessed independently in Add Health, making it impossible to fully enumerate Hispanic/Latino participants that also identify as Multiracial. This is because in contrast to the current federal racial/ethnic categorization schema, some (but not all) Hispanic/Latino individuals regard this label as a racial identity.^23,45^

### Measures

Because differential exposure to ACEs may be linked to racial disparities in health through the stress pathway, we investigated physical, mental, and behavioral outcomes (groupings described below) for which (1) Multiracial Americans experience relatively poor health outcomes, and (2) are associated with ACEs and overactivation of the stress pathway.^46–50^ We measured outcomes once for each individual in either Wave 3 (ages 18-26) or 4 (24-32) based on data availability. Appendix A contains details on construction of outcomes and covariates.

- *Physical health: metabolic syndrome (MetS), hypertension, and asthma*. We assessed MetS using clinically-relevant cutoffs for eight biomarkers across five categories: (1) hypertension (systolic blood pressure ≥ 130 mmHg, diastolic blood pressure ≥ 80 mmHg),^51^ self-reported ever diagnosed with hypertension, or self-reported current hypertension medication use), (2) waist circumference (> 102 cm for males, > 80 cm for females), (3) triglycerides (top 3 deciles for males, top 2 for females), (4) high-density lipoprotein (bottom 2 deciles for males, bottom 3 for females), and (5) diabetes (glycated hemoglobin > 5.7%, self-reported anti-diabetic medication use, or self-reported diabetes diagnosis).^52^ We categorized individuals with values exceeding cutoffs in three or more of the five categories as having MetS.^52–56^ Hypertension coding when considering it as a separate outcome mirrored that used for the MetS measure. We coded participants as having asthma if they indicated a previous diagnosis.^57^
- *Mental health: depression and anxiety*. We classified participants as having depression if they indicated using prescription depression medications in the last year or having a previous depression diagnosis, and as having anxiety if they indicated a previous anxiety diagnosis.
- *Behavioral health: suicidal ideation and drug use*. We coded participants as having suicidal ideation if they indicated seriously thinking about committing suicide in the last year, and using drugs if they indicated ever using cocaine, crystal meth, heroin, other illegal drugs other than marijuana, or ever misusing prescription drugs.

The original ACEs questionnaire included ten questions each covering a domain of adverse experiences: emotional, physical, and sexual abuse, emotional and physical neglect, parental separation or divorce, mother treated violently, household substance abuse, household mental illness, and incarceration of household member.^58^ Researchers constructing ACEs scores using Add Health data have used varying sets of questions about events occurring before age 18 to approximate the original ACEs questionnaire.^59–63^ We used a modified version of the widest set of questions available, some of which were answered by participants’ parents in Wave 1, and others retrospectively by participants during Waves 3 or 4. We dichotomized and summed responses to create a summary score from zero to ten. We categorized participants reporting four or more ACEs as exposed, as a previous meta-analysis found elevated risk above this level for all studied negative physical, mental, and behavioral health outcomes.^64^ We conducted sensitivity analyses using the summary score instead of a dichotomized exposure. Questions and response categories used are shown in Appendix B.

We identified confounders using a directed acyclic graph. Our models were adjusted for age, sex, parental education, household size-adjusted income, parental support, and neighborhood disadvantage score as confounders. We drew age and sex from Wave 4 to match timing of collected biomarkers. The Wave 4 questionnaire asked respondents to report their gender but only provided “male” and “female” as response options; we thus consider this variable to represent biological sex. Following previous Add Health analyses, we categorized parental education as the higher of either parent’s Wave 1 parental education after categorization into a five-level ordinal variable (less than high school, completed vocational school, or GED; high school diploma; some college; college graduate or greater).^65^ We calculated household size-adjusted income as total reported pre-tax household income divided by the square root of total reported household members at Wave 1.^66^ We coded parental support as the mean of Likert-scale responses (very much, quite a bit, some, a little, none at all) to five Wave 1 questions about participants’ relationship and communication with their parents.^67,68^ We constructed a crude neighborhood disadvantage score by averaging five census tract-level proportions from participants’ Wave 1 residence: (1) percent households with incomes below the federal poverty level, (2) percent households receiving public assistance, (3) civilian unemployment rate, (4) percent persons 25 years or older with no high school diploma or equivalency, and (5) percent female-headed households.^55,69^

### Statistical analyses

For each outcome, we specified a modified Poisson model to estimate risk ratios.^70^ We visually assessed assumptions of linearity between exposure and log-transformed risk of outcomes for each racial group by comparing observed data with linear model-generated smooth plots. All models interacted ACEs and race to estimate subgroup-specific associations. Because our aim was to understand if associations are stronger for Multiracial people, we specified Multiracial participants as the reference group in all interactions. We also conducted sensitivity analyses with Whites as the referent group to compare findings with previous studies which have used this approach.

We used complex survey weights corresponding to a cross-sectional multi-wave analysis to produce nationally representative estimates using the “survey” package in R.^71,72^ Because 55% of observations were missing data on an ACE component, 21% on a covariate, and 40% on an outcome (frequencies presented in Appendix C), we used multiple imputation including all outcomes in imputations models, and pooled results across 20 imputed datasets.^73^ We imputed data using the “mi” and “mitools” packages in R.^74,75^ We summed and dichotomized ACE scores in regression analyses after imputation of missing ACE component variables.

We assessed interactions on the relative scale by estimating subgroup specific risk ratios (RRs), and on the absolute scale by estimating the interaction contrast, scaled up to reflect excess cases per 1,000 (formula and details in Appendix D).^76^ We assessed statistical differences between group RRs using p-values with pre-specified α = 0.05 from global Wald tests for significance of interaction terms. We deemed excess case estimates with 95% confidence intervals (CI) not containing zero (null value) as significant.

The UC Berkeley Office for Protection of Human Subjects determined that this study did not meet the threshold definition of human subjects research; we conducted analyses in 2023.

## Results

Post-hoc analysis led us to exclude individuals who identified as “Other” race alone due to small sample size (n=23) and overly wide CIs. Table 1 shows key characteristics (unweighted counts and weighted statistics pooled from imputations) in the overall study sample. Sample size in models with self-reported outcome data was 12,372. Biomarkers can be affected by pregnancy, thus we excluded 445 pregnant participants from the MetS and hypertension models. In the overall study sample, there were 7,742 (74%) White, 2,915 (17%) Black, 805 (3.2%) Asian, 76 (0.6%) AI/NA, and 834 (5.8%) Multiracial participants. Mean age, sex ratios, and parental support scores were approximately even across racial groups. White and Asian participants’ households had parents with higher educational attainment, higher equivalence-scaled income, and lower neighborhood disadvantage scores compared to Black and AI/NA participants. On these measures, Multiracial participants were less advantaged than White and Asian but more advantaged than Black and AI/NA participants.

**Table 1.** Participant characteristics and outcomes^a^ stratified by race, Add Health 1994-2008

| **Characteristic** | **Overall**<br>12,372 (100%) | **White**<br>7,742 (74%) | **Black**<br>2,915 (17%) | **Asian**<br>805 (3.2%) | **AI/NA**<br>76 (0.6%) | **Multiracial**<br>834 (5.8%) |
| --- | --- | --- | --- | --- | --- | --- |
| Male sex | 5,778 (51%) | 3,672 (51%) | 1,267 (50%) | 418 (53%) | 41 (64%) | 380 (50%) |
| Age | 29.0 | 28.9 | 29.2 | 29.2 | 28.8 | 28.8 |
| Highest parental education |  |  |  |  |  |  |
| Less than high school<sup>b</sup> | 1,187 (11%) | 685 (9.6%) | 356 (17%) | 69 (13%) | 13 (29%) | 64 (11%) |
| High school diploma | 3,883 (35%) | 2,520 (34%) | 959 (42%) | 164 (24%) | 19 (34%) | 221 (33%) |
| Some college | 2,221 (18%) | 1,405 (19%) | 500 (16%) | 106 (10%) | 19 (24%) | 191 (22%) |
| College graduate or greater | 4,897 (36%) | 3,039 (38%) | 1,043 (26%) | 445 (53%) | 24 (14%) | 346 (34%) |
| Household income<sup>c</sup> | 23.5 | 25.5 | 14.4 | 27.2 | 13.2 | 21.2 |
| Parental support index (0-5) | 3.33 | 3.35 | 3.26 | 3.30 | 3.38 | 3.30 |
| Neighborhood disadvantage (0-1) | 0.14 | 0.12 | 0.23 | 0.13 | 0.23 | 0.15 |
| Elevated ($\geq 4$) ACEs<sup>d</sup> | 4,402 (27%) | 3,207 (25%) | 648 (33%) | 260 (21%) | 15 (40%) | 272 (35%) |
| MetS | 3,131 (29%) | 1,799 (27%) | 852 (38%) | 221 (27%) | 35 (56%) | 224 (31%) |
| Hypertension | 6,082 (52%) | 3,775 (51%) | 1,464 (55%) | 390 (46%) | 46 (66%) | 407 (50%) |
| Depression | 2,195 (33%) | 1,652 (35%) | 296 (26%) | 53 (17%) | 11 (32%) | 183 (35%) |
| Asthma | 1,867 (15%) | 1,134 (15%) | 453 (15%) | 90 (10%) | 14 (16%) | 176 (24%) |
| Anxiety | 1,518 (13%) | 1,179 (15%) | 182 (6.0%) | 26 (2.9%) | 4 (3.3%) | 127 (18%) |
| Suicidal ideation | 845 (7.4%) | 527 (7.2%) | 188 (7.4%) | 48 (6.0%) | 8 (15%) | 74 (10%) |
| Drug use | 3,965 (36%) | 3,049 (41%) | 356 (12%) | 197 (25%) | 30 (46%) | 333 (42%) |

Abbreviations: ACE = adverse childhood experience; MetS = metabolic syndrome
<sup>a</sup> Counts are unweighted; proportions (for categorical variables) and means (for continuous) are pooled estimates from 20 survey-weighted imputations
<sup>b</sup> Includes completed vocational school or GED
<sup>c</sup> Equivalence-scaled to adjust for household size
<sup>d</sup> ACE components measured variously across Waves 1, 3, and 4, and summed after imputation; see Appendix A for details

Exposure to elevated ACEs was highest among AI/NA (40%), Multiracial (35%), and Black participants (33%). Multiracial participants had the highest prevalence of asthma (24%) and anxiety (18%), and along with Whites, the highest prevalence of depression (35%). For all other outcomes, AI/NA participants had the highest prevalence. Asian participants reported the lowest prevalence of all outcomes except for drug use, for which Black participants had the lowest prevalence (12%). White and Asian participants had the lowest prevalence of MetS (27%).

Table 2 summarizes within-group RRs and 95% CIs for each outcome. For the overall sample, increased risks associated with elevated ACEs were null for MetS and hypertension, small (4-5%) for asthma, anxiety, and suicidal ideation, and stronger (10-13%) for depression and drug use. Risks of asthma, anxiety, depression, and drug use increased 12-14% among Multiracial participants, and of drug use by 30% for AI/NA participants. Associations were strongest with depression and drug use (8-13% increased risk) for White and Black participants, and with depression, suicidal ideation, and drug use (8-10%) for Asians. We estimated RRs for interaction using Multiracial participants as the referent group; Table 3 summarizes the exponentiated interaction term betas from each model. Values covered by CIs around interaction terms in the asthma model were also consistent with weaker associations for White (0.92, 95% CI: 0.85, 1.00), Black (0.91, 95% CI: 0.83, 1.00) and Asian (0.89, 95% CI: 0.79, 1.00) participants. CIs around interaction terms in the anxiety model were also consistent with stronger association for Multiracial individuals compared to all other groups. CIs for (Black (0.93, 95% CI: 0.86, 0.99), Asian (0.87, 95% CI: 0.82, 0.94), AI/NA (0.89, 95% CI: 0.81, 0.97) participants did not include the null. Global Wald tests for interaction were only significant for anxiety (p < 0.001).

**Table 2.** Adjusted^a^ overall and within-group RRs and 95% CIs associated with elevated ACEs, Add Health 1994-2008

| **Outcome** | **Overall** | **White** | **Black** | **Asian** | **AI/NA** | **Multiracial** |
| --- | --- | --- | --- | --- | --- | --- |
| MetS | 1.00 (0.98, 1.02) | 1.03 (0.94, 1.12) | 1.00 (0.97, 1.02) | 1.00 (0.96, 1.05) | 0.99 (0.87, 1.13) | 1.00 (0.78, 1.28) |
| Hypertension | 0.99 (0.97, 1.01) | 0.99 (0.97, 1.01) | 1.01 (0.96, 1.05) | 1.02 (0.90, 1.16) | 0.86 (0.69, 1.07) | 0.98 (0.91, 1.04) |
| Asthma | **1.04 (1.03, 1.06)** | **1.04 (1.02, 1.06)** | 1.02 (0.98, 1.07) | 1.00 (0.93, 1.08) | 1.15 (0.93, 1.42) | **1.13 (1.04, 1.22)** |
| Depression | **1.13 (1.10, 1.16)** | **1.13 (1.09, 1.16)** | **1.13 (1.08, 1.18)** | 1.10 (0.98, 1.24) | 1.21 (0.90, 1.61) | **1.14 (1.07, 1.22)** |
| Anxiety | **1.05 (1.03, 1.07)** | **1.05 (1.03, 1.08)** | 1.03 (1.00, 1.06) | 0.98 (0.95, 1.00) | 0.99 (0.92, 1.07) | **1.12 (1.04, 1.19)** |
| Suicidal ideation | **1.05 (1.03, 1.07)** | **1.05 (1.03, 1.07)** | **1.05 (1.01, 1.09)** | 1.08 (1.00, 1.17) | 1.07 (0.84, 1.37) | 1.05 (0.98, 1.12) |
| Drug use | **1.10 (1.08, 1.13)** | **1.11 (1.08, 1.13)** | **1.08 (1.03, 1.12)** | 1.08 (0.95, 1.23) | **1.30 (1.06, 1.60)** | **1.13 (1.07, 1.20)** |

Abbreviations: RR = risk ratio, CI = confidence interval, ACE = adverse childhood experience; MetS = metabolic syndrome
Boldface indicates statistical significance (95% CI does not include null)
<sup>a</sup> Models adjusted for participant age, sex, parental education, household size-adjusted income, parental support, and neighborhood disadvantage score

**Table 3.** Exponentiated interaction^a^ term betas and 95% CIs from regression analyses, Add Health 1994-2008

| Outcome | White | Black | Asian | AI/NA | Multiracial | p-value |
| --- | --- | --- | --- | --- | --- | --- |
| MetS | 0.97 (0.89, 1.06) | 0.98 (0.89, 1.07) | 0.97 (0.82, 1.14) | 0.97 (0.76, 1.24) | (ref.) | 0.98 |
| Hypertension | 1.01 (0.94, 1.09) | 1.03 (0.95, 1.12) | 1.05 (0.91, 1.21) | 0.88 (0.70, 1.10) | (ref.) | 0.65 |
| Asthma | 0.92 (0.85, 1.00) | 0.91 (0.83, 1.00) | 0.89 (0.79, 1.00) | 1.02 (0.82, 1.28) | (ref.) | 0.21 |
| Depression | 0.98 (0.91, 1.06) | 0.99 (0.91, 1.07) | 0.96 (0.84, 1.10) | 1.05 (0.78, 1.42) | (ref.) | 0.97 |
| Anxiety | 0.94 (0.88, 1.02) | **0.93 (0.86, 0.99)** | **0.87 (0.82, 0.94)** | **0.89 (0.81, 0.97)** | (ref.) | *** < **0.001** |
| Suicidal ideation | 1.00 (0.93, 1.08) | 1.00 (0.92, 1.09) | 1.03 (0.93, 1.15) | 1.02 (0.79, 1.32) | (ref.) | 0.96 |
| Drug use | 0.98 (0.92, 1.04) | 0.95 (0.89, 1.02) | 0.96 (0.83, 1.11) | 1.15 (0.93, 1.42) | (ref.) | 0.29 |

Abbreviations: CI = confidence interval, ACE = adverse childhood experience; MetS = metabolic syndrome
Boldface indicates statistical significance (global Wald test \* $p < 0.05$ , \*\* $p < 0.01$ , \*\*\* $p < 0.001$ )
<sup>a</sup> Race × elevated ACEs interaction, Multiracial as referent group

Table 4 summarizes excess cases per 1,000 associated with elevated ACEs, with the Multiracial group as the reference. Results were mostly consistent with interaction RRs in Table 3. CIs around estimates in the asthma model suggested significantly weaker fewer excess cases for White (−123, 95% CI: -251, -4), Black (−141, 95% CI: -285, -6), and Asian (−169, 95% CI: -334, - 7) participants. There were significantly fewer excess cases of anxiety for Black (−100, 95% CI: - 189, -10), Asian (−163, 95% CI: -247, -79), and AI/NA (−144, 95% CI: -252, -42) participants; the estimate and CI for Whites (−71, 95% CI: -165, 25) also suggested a weaker association compared to Multiracial participants. CIs around excess case estimates were wide for some outcomes, especially for the AI/NA and Asian groups.

**Table 4.**
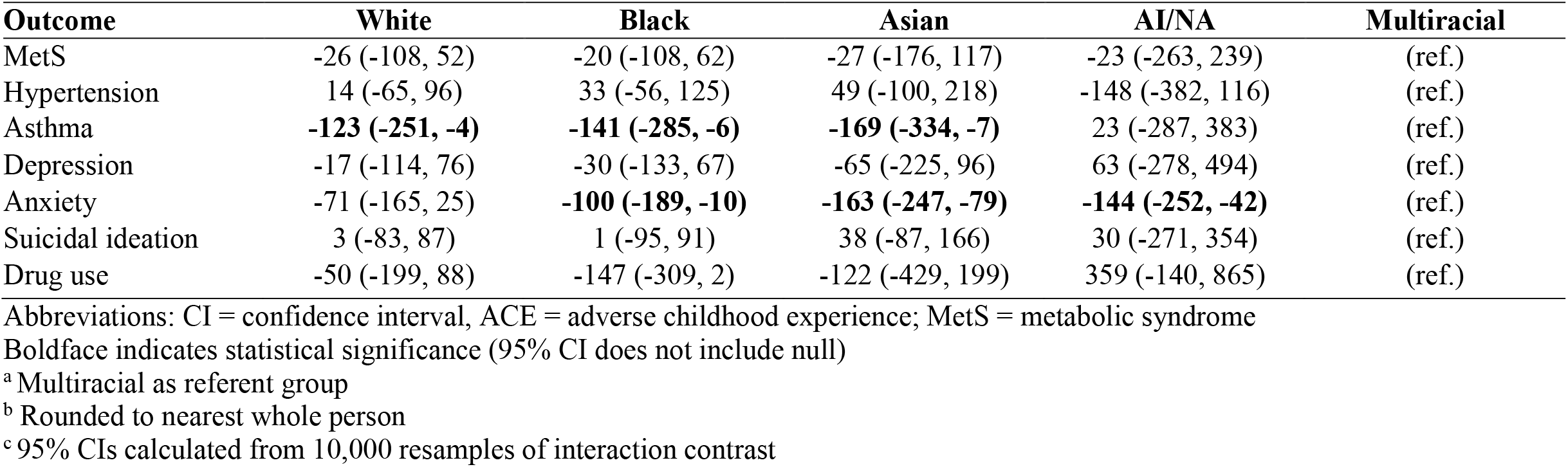
Excess^a^ cases^b^ per 1,000 and 95% CIs^c^ associated with elevated ACEs, Add Health 1994-2008

Appendix E summarizes results from the first sensitivity analysis which specified summary ACE score (rather than ≥ 4 ACEs) as the exposure, keeping Multiracial participants as the reference. Direction and significance of results were similar to the main analysis. Appendix F displays results from the second sensitivity analysis which specified White participants as the reference, keeping elevated ACEs as the exposure. Directions of results were consistent with the main analysis.

## Discussion

Our study aimed to explore whether the association of ACEs with physical, mental, and behavioral health outcomes differs by race, and in particular, if Multiracial individuals should be prioritized for interventions given their unique ACEs and health risk profile. We found evidence that, compared to Black, Asian, and AI/NA participants, Multiracial people experience greater excess cases of anxiety (and relative strength of association) associated with ACEs, regardless of ACEs coding scheme. We also estimated that Multiracial participants have more excess cases of asthma associated with elevated ACEs compared to most other racial groups examined.

Data from the National Health Interview Survey show that prevalence of asthma and anxious symptoms are disproportionately and significantly higher among Multiracial people than other racial groups.^31,33^ ACEs may thus contribute meaningfully to the disparity in asthma and anxiety for Multiracial adults. Future studies that estimate attributable risk will aid our understanding of the potential health equity benefits of preventing exposure to ACEs.

Although ACEs are a well-established risk factor for a variety of poor adulthood health outcomes, the structural and social processes that determine disparities in exposure to ACEs, and the causes of variability in race-specific associations are not well understood. Despite Multiracial children’s high rate of exposure to ACEs, and even though the 2020 Census reports that one in ten people selected two or more races^77^, health equity research continues to overlook the Multiracial population. While our study does not explore structural processes or the causes of disparities in exposure and associations, it does highlight the contribution of ACEs to the burden of respiratory and mental health morbidity among Multiracial adults.

## Limitations

Our study had strengths, including a uniquely large representative and longitudinal sample of Multiracial participants and multiple investigated outcomes. However, there were also limitations. First, the racial categories used in this analysis and health research generally collapse large amounts of within-group variability in patterns of health and disadvantage; subgroup analyses thus produce only a rough proxy for markers of elevated risk and should be contextualized with other knowledge. Second, our ACEs measure is unique to Add Health and had high levels of missingness. Measures for asthma, anxiety, and depression were self-reported, and may be biased downward for racial groups with less healthcare access or where mental health conditions are especially stigmatized.^78^ Our measure of parental education used highest reported level between either parent rather than primary caregiver, which some research suggests may better predict child health.^79^ Finally, small sample sizes particularly for Asian and AI/NA groups contributed to wide CIs around excess case estimates; future studies should repeat this analysis in larger longitudinal cohorts with greater numbers of Asian and AI/NA participants.

## Conclusion

To our knowledge, this is the first study finding that exposure to ACEs is associated with excess risk of anxiety and asthma for Multiracial people, a finding that would be obscured if Multiracial people were recategorized into an “other” or monoracial categories. Given the large and growing size of the Multiracial population and inequitable rates of exposure to ACEs among Multiracial children, addressing and preventing ACEs is an urgent health equity issue. While intervening to reduce exposure to ACEs will be universally beneficial for all racial groups, such programs may especially benefit the respiratory and mental health of the Multiracial population. Future studies should continue to examine the aggregate and group-specific population health benefits of preventing ACEs to improve health equity.

## Data Availability

This analysis used restricted data (made available through a data use agreement); a public-use subset is available online.

https://addhealth.cpc.unc.edu/data/

## Acknowledgements

The authors would like to thank the Add Health study participants for donating their time and information to the study of social experiences and health. Thanks as well to Kim Harley for facilitating access to the data, to Christian Jackson for reviewing the code used in this analysis, and to David Rehkopf for thought partnership during manuscript development.

## References

1. Centers for Disease Control and Prevention. Adverse Childhood Experiences (ACEs). Published April 2, 2021. Accessed April 5, 2022. https://www.cdc.gov/violenceprevention/aces/index.html

2. Petruccelli K, Davis J, Berman T. Adverse childhood experiences and associated health outcomes: A systematic review and meta-analysis. Child Abuse Negl. 2019;97:104127. doi:10.1016/j.chiabu.2019.104127

3. Kalmakis KA, Chandler GE. Health consequences of adverse childhood experiences: A systematic review. J Am Assoc Nurse Pract. 2015;27(8):457–465. doi:10.1002/2327-6924.12215

4. Merrick MT, Ford DC, Ports KA, Guinn AS. Prevalence of Adverse Childhood Experiences From the 2011-2014 Behavioral Risk Factor Surveillance System in 23 States. JAMA Pediatr. 2018;172(11):1038. doi:10.1001/jamapediatrics.2018.2537

5. Felitti VJ, Anda RF, Nordenberg D, et al. Relationship of Childhood Abuse and Household Dysfunction to Many of the Leading Causes of Death in Adults. Am J Prev Med. 1998;14(4):245–258. doi:10.1016/S0749-3797(98)00017-8

6. Jones TM, Nurius P, Song C, Fleming CM. Modeling life course pathways from adverse childhood experiences to adult mental health. Child Abuse Negl. 2018;80:32–40. doi:10.1016/j.chiabu.2018.03.005

7. Nurius PS, Green S, Logan-Greene P, Borja S. Life course pathways of adverse childhood experiences toward adult psychological well-being: A stress process analysis. Child Abuse Negl. 2015;45:143–153. doi:10.1016/j.chiabu.2015.03.008

8. Manyema M, Norris SA, Richter LM. Stress begets stress: the association of adverse childhood experiences with psychological distress in the presence of adult life stress. BMC Public Health. 2018;18(1):835. doi:10.1186/s12889-018-5767-0

9. Hertzman C, Power C. Health and Human Development: Understandings From Life-Course Research. Dev Neuropsychol. 2003;24(2-3):719–744. doi:10.1080/87565641.2003.9651917

10. Stewart JA. The Detrimental Effects of Allostasis: Allostatic Load as a Measure of Cumulative Stress. J Physiol Anthropol. 2006;25(1):133–145. doi:10.2114/jpa2.25.133

11. Kelly-Irving M, Mabile L, Grosclaude P, Lang T, Delpierre C. The embodiment of adverse childhood experiences and cancer development: potential biological mechanisms and pathways across the life course. Int J Public Health. 2013;58(1):3–11. doi:10.1007/s00038-012-0370-0

12. Labonté B, Suderman M, Maussion G, et al. Genome-wide Epigenetic Regulation by Early-Life Trauma. Arch Gen Psychiatry. 2012;69(7). doi:10.1001/archgenpsychiatry.2011.2287

13. Moore LD, L. T, Fan G. DNA Methylation and Its Basic Function. Neuropsychopharmacology. 2013;38(1):23–38. doi:10.1038/npp.2012.112

14. CDC. What is Epigenetics? | CDC. Centers for Disease Control and Prevention. Published August 3, 2020. Accessed April 26, 2022. https://www.cdc.gov/genomics/disease/epigenetics.htm

15. Wiss DA, Brewerton TD. Adverse Childhood Experiences and Adult Obesity: A Systematic Review of Plausible Mechanisms and Meta-Analysis of Cross-Sectional Studies. Physiol Behav. 2020;223:112964. doi:10.1016/j.physbeh.2020.112964

16. Su S, Jimenez MP, Roberts CTF, Loucks EB. The Role of Adverse Childhood Experiences in Cardiovascular Disease Risk: a Review with Emphasis on Plausible Mechanisms. Curr Cardiol Rep. 2015;17(10):88. doi:10.1007/s11886-015-0645-1

17. Vig KD, Paluszek MM, Asmundson GJG. ACEs and physical health outcomes. In: Adverse Childhood Experiences. Elsevier; 2020:71–90. doi:10.1016/B978-0-12-816065-7.00005-7

18. Wekerle C, Hébert M, Daigneault I, Fortin-Langelier E, Smith S. ACEs, sexual violence, and sexual health. In: Adverse Childhood Experiences. Elsevier; 2020:91–118. doi:10.1016/B978-0-12-816065-7.00006-9

19. Giano Z, Wheeler DL, Hubach RD. The frequencies and disparities of adverse childhood experiences in the U.S. BMC Public Health. 2020;20(1):1327. doi:10.1186/s12889-020-09411-z

20. Skewes MC, Blume AW. Understanding the link between racial trauma and substance use among American Indians. Am Psychol. 2019;74(1):88–100. doi:10.1037/amp0000331

21. Kenney MK, Singh GK. Adverse Childhood Experiences among American Indian/Alaska Native Children: The 2011-2012 National Survey of Children’s Health. Scientifica. 2016;2016:1–14. doi:10.1155/2016/7424239

22. Cronholm PF, Forke CM, Wade R, et al. Adverse Childhood Experiences. Am J Prev Med. 2015;49(3):354–361. doi:10.1016/j.amepre.2015.02.001

23. Udry JR, Li RM, Hendrickson-Smith J. Health and Behavior Risks of Adolescents with Mixed-Race Identity. Am J Public Health. 2003;93(11):1865–1870. doi:10.2105/AJPH.93.11.1865

24. Straka BC, Gaither SE, Acheson SK, Swartzwelder HS. “Mixed” Drinking Motivations: A Comparison of Majority, Multiracial, and Minority College Students. Soc Psychol Personal Sci. Published online November 19, 2019:194855061988329. doi:10.1177/1948550619883294

25. Pang YC. The Relationship between Perceived Discrimination, Economic Pressure, Depressive Symptoms, and Educational Attainment of Ethnic Minority Emerging Adults: The Moderating Role of School Connectedness during Adolescence.Master of Science. Iowa State University, Digital Repository; 2015. doi:10.31274/etd-180810-4045

26. Doyle JM. THE PHYSICAL AND MENTAL HEALTH OF MULTIRACIAL ADOLESCENTS IN THE UNITED STATES. University of Michigan; 2007.

27. Goodhines PA, Desalu JM, Zaso MJ, Gellis LA, Park A. Sleep Problems and Drinking Frequency among Urban Multiracial and Monoracial Adolescents: Role of Discrimination Experiences and Negative Mood. J Youth Adolesc. 2020;49(10):2109–2123. doi:10.1007/s10964-020-01310-1

28. Choi Y, Harachi TW, Gillmore MR, Catalano RF. Are multiracial adolescents at greater risk? Comparisons of rates, patterns, and correlates of substance use and violence between monoracial and multiracial adolescents. Am J Orthopsychiatry. 2006;76(1):86–97. doi:10.1037/0002-9432.76.1.86

29. Goings TC, Salas-Wright CP, Howard MO, Vaughn MG. Substance use among bi/multiracial youth in the United States: Profiles of psychosocial risk and protection. Am J Drug Alcohol Abuse. 2018;44(2):206–214. doi:10.1080/00952990.2017.1359617

30. National Center for Health Statistics. Summary Health Statistics, Selected Circulatory Diseases: National Health Interview Survey, 2018. Published 2018. Accessed May 16, 2023. https://ftp.cdc.gov/pub/Health_Statistics/NCHS/NHIS/SHS/2018_SHS_Table_A-1.pdf

31. National Center for Health Statistics. Summary Health Statistics, Respiratory Diseases: National Health Interview Survey, 2018. Published 2018. Accessed May 16, 2023. https://ftp.cdc.gov/pub/Health_Statistics/NCHS/NHIS/SHS/2018_SHS_Table_A-2.pdf

32. National Center for Health Statistics. Summary Health Statistics, Body Mass Index: National Health Interview Survey, 2018. Published 2018. Accessed May 16, 2023. https://ftp.cdc.gov/pub/Health_Statistics/NCHS/NHIS/SHS/2018_SHS_Table_A-15.pdf

33. National Center for Health Statistics. Summary Health Statistics, Mental Health Conditions: National Health Interview Survey, 2018.Published 2018. Accessed March 14, 2023. https://ftp.cdc.gov/pub/Health_Statistics/NCHS/NHIS/SHS/2018_SHS_Table_A-8.pdf

34. Summary Health Statistics: National Health Interview Survey. Centers for Disease Control and Prevention; 2018. Accessed October 6, 2020. https://ftp.cdc.gov/pub/Health_Statistics/NCHS/NHIS/SHS/2018_SHS_Table_A-7.pdf

35. Subica AM, Agarwal N, Sullivan JG, Link BG. Obesity and Associated Health Disparities Among Understudied Multiracial, Pacific Islander, and American Indian Adults: Understudied Racial Disparities in Obesity. Obesity. 2017;25(12):2128–2136. doi:10.1002/oby.21954

36. Charmaraman L, Woo M, Quach A, Erkut S. How have researchers studied multiracial populations? A content and methodological review of 20 years of research. Cultur Divers Ethnic Minor Psychol. 2014;20(3):336–352. doi:10.1037/a0035437

37. Facente SN, Lam-Hine T, Bhatta DN, Hecht J. Impact of Racial Categorization on Effect Estimates: An HIV Stigma Analysis. Am J Epidemiol. Published online January 5, 2022:kwab289. doi:10.1093/aje/kwab289

38. Hall T, Rooks R, Kaufman C. Intersections of Adverse Childhood Experiences, Race and Ethnicity and Asthma Outcomes: Findings from the Behavioral Risk Factor Surveillance System. Int J Environ Res Public Health. 2020;17(21):8236. doi:10.3390/ijerph17218236

39. LaBrenz CA, O’Gara JL, Panisch LS, Baiden P, Larkin H. Adverse childhood experiences and mental and physical health disparities: the moderating effect of race and implications for social work. Soc Work Health Care. 2020;59(8):588–614. doi:10.1080/00981389.2020.1823547

40. Youssef NA, Belew D, Hao G, et al. Racial/ethnic differences in the association of childhood adversities with depression and the role of resilience. J Affect Disord. 2017;208:577–581. doi:10.1016/j.jad.2016.10.024

41. Stinson EA, Sullivan RM, Peteet BJ, et al. Longitudinal Impact of Childhood Adversity on Early Adolescent Mental Health During the COVID-19 Pandemic in the ABCD Study Cohort: Does Race or Ethnicity Moderate Findings? Biol Psychiatry Glob Open Sci. 2021;1(4):324–335. doi:10.1016/j.bpsgos.2021.08.007

42. Assini-Meytin LC, Fix RL, Green KM, Nair R, Letourneau EJ. Adverse Childhood Experiences, Mental Health, and Risk Behaviors in Adulthood: Exploring Sex, Racial, and Ethnic Group Differences in a Nationally Representative Sample. J Child Adolesc Trauma. 2022;15(3):833–845. doi:10.1007/s40653-021-00424-3

43. Lee RD, Chen J. Adverse childhood experiences, mental health, and excessive alcohol use: Examination of race/ethnicity and sex differences. Child Abuse Negl. 2017;69:40–48. doi:10.1016/j.chiabu.2017.04.004

44. Harris KM. The Add Health Study: Design and Accomplishments. Published online 2013. doi:10.17615/C6TW87

45. Taylor P, Lopez MH, Martínez J, Velasco G. When Labels Don’t Fit: Hispanics and Their Views of Identity. Pew Research Center’s Hispanic Trends Project. Published April 4, 2012. Accessed July 22, 2022. https://www.pewresearch.org/hispanic/2012/04/04/when-labels-dont-fit-hispanics-and-their-views-of-identity/

46. Williams DR, Neighbors H. Racism, discrimination and hypertension: evidence and needed research. Ethn Dis. 2001;11(4):800–816.

47. National Heart, Lung, and Blood Institute. Metabolic Syndrome | NHLBI, NIH. Published December 2020. Accessed March 3, 2021. https://www.nhlbi.nih.gov/health-topics/metabolic-syndrome

48. Cuevas AG, Williams DR, Albert MA. Psychosocial Factors and Hypertension. Cardiol Clin. 2017;35(2):223–230. doi:10.1016/j.ccl.2016.12.004

49. Exley D, Norman A, Hyland M. Adverse childhood experience and asthma onset: a systematic review. Eur Respir Rev. 2015;24(136):299–305. doi:10.1183/16000617.00004114

50. Chen E, Miller GE. Stress and inflammation in exacerbations of asthma. Brain Behav Immun. 2007;21(8):993–999. doi:10.1016/j.bbi.2007.03.009

51. Whelton PK, Carey RM, Aronow WS, et al. 2017 ACC/AHA/AAPA/ABC/ACPM/AGS/APhA/ASH/ASPC/NMA/PCNA Guideline for the Prevention, Detection, Evaluation, and Management of High Blood Pressure in Adults: A Report of the American College of Cardiology/American Heart Association Task Force on Clinical Practice Guidelines. Hypertension. 2018;71(6):e13–e115. doi:10.1161/HYP.0000000000000065

52. Bohr AD, Laurson K, McQueen MB. A novel cutoff for the waist-to-height ratio predicting metabolic syndrome in young American adults. BMC Public Health. 2016;16(1):295. doi:10.1186/s12889-016-2964-6

53. Gaydosh L, Schorpp KM, Chen E, Miller GE, Harris KM. College completion predicts lower depression but higher metabolic syndrome among disadvantaged minorities in young adulthood. Proc Natl Acad Sci. 2018;115(1):109–114. doi:10.1073/pnas.1714616114

54. Miller GE, Chen E, Yu T, Brody GH. Youth Who Achieve Upward Socioeconomic Mobility Display Lower Psychological Distress But Higher Metabolic Syndrome Rates as Adults: Prospective Evidence From Add Health and MIDUS. J Am Heart Assoc. 2020;9(9). doi:10.1161/JAHA.119.015698

55. Martin CL, Kane JB, Miles GL, Aiello AE, Harris KM. Neighborhood disadvantage across the transition from adolescence to adulthood and risk of metabolic syndrome. Health Place. 2019;57:131–138. doi:10.1016/j.healthplace.2019.03.002

56. Colen CG, Pinchak NP, Barnett KS. Racial Disparities in Health Among College-Educated African Americans: Can Attendance at Historically Black Colleges or Universities Reduce the Risk of Metabolic Syndrome in Midlife? Am J Epidemiol. Published online November 5, 2020:kwaa245. doi:10.1093/aje/kwaa245

57. Harris KM, Gordon-Larsen P, Chantala K, Udry JR. Longitudinal Trends in Race/Ethnic Disparities in Leading Health Indicators From Adolescence to Young Adulthood. Arch Pediatr Adolesc Med. 2006;160(1):74. doi:10.1001/archpedi.160.1.74

58. About the CDC-Kaiser ACE Study |Violence Prevention|Injury Center|CDC. Published November 3, 2021. Accessed January 10, 2022. https://www.cdc.gov/violenceprevention/aces/about.html

59. Easterlin MC, Chung PJ, Leng M, Dudovitz R. Association of Team Sports Participation With Long-term Mental Health Outcomes Among Individuals Exposed to Adverse Childhood Experiences. JAMA Pediatr. 2019;173(7):681. doi:10.1001/jamapediatrics.2019.1212

60. LeTendre ML, Reed MB. The Effect of Adverse Childhood Experience on Clinical Diagnosis of a Substance Use Disorder: Results of a Nationally Representative Study. Subst Use Misuse. 2017;52(6):689–697. doi:10.1080/10826084.2016.1253746

61. Brumley LD, Jaffee SR, Brumley BP. Pathways from Childhood Adversity to Problem Behaviors in Young Adulthood: The Mediating Role of Adolescents’ Future Expectations. J Youth Adolesc. 2017;46(1):1–14. doi:10.1007/s10964-016-0597-9

62. Lee H, Kim Y, Terry J. Adverse childhood experiences (ACEs) on mental disorders in young adulthood: Latent classes and community violence exposure. Prev Med. 2020;134:106039. doi:10.1016/j.ypmed.2020.106039

63. Otero C. Adverse Childhood Experiences (ACEs) and Timely Bachelor’s Degree Attainment. Soc Sci. 2021;10(2):44. doi:10.3390/socsci10020044

64. Hughes K, Bellis MA, Hardcastle KA, et al. The effect of multiple adverse childhood experiences on health: a systematic review and meta-analysis. Lancet Public Health. 2017;2(8):e356–e366. doi:10.1016/S2468-2667(17)30118-4

65. Goodman E, Slap GB, Huang B. The Public Health Impact of Socioeconomic Status on Adolescent Depression and Obesity. Am J Public Health. 2003;93(11):1844–1850. doi:10.2105/AJPH.93.11.1844

66. Organization for Economic Cooperation and Development. Adjusting Household Incomes: Equivalence Scales. Organization for Economic Cooperation and Development Accessed April 28, 2022. https://www.oecd.org/els/soc/OECD-Note-EquivalenceScales.pdf

67. Chen P, Harris KM. Association of Positive Family Relationships With Mental Health Trajectories From Adolescence to Midlife. JAMA Pediatr. 2019;173(12):e193336. doi:10.1001/jamapediatrics.2019.3336

68. Sieving RE, McNeely CS, Blum RWm. Maternal Expectations, Mother-Child Connectedness, and Adolescent Sexual Debut. Arch Pediatr Adolesc Med. 2000;154(8):809. doi:10.1001/archpedi.154.8.809

69. Ross CE, Mirowsky J. Neighborhood disadvantage, disorder, and health. J Health Soc Behav. 2001;42(3):258–276.

70. Zou G. A Modified Poisson Regression Approach to Prospective Studies with Binary Data. Am J Epidemiol. 2004;159(7):702–706. doi:10.1093/aje/kwh090

71. Lumley T. Analysis of Complex Survey Samples. J Stat Softw. 2004;9(8). doi:10.18637/jss.v009.i08

72. Chen P. Guidelines for Analyzing Add Health Data. Published online 2014. doi:10.17615/C6BW8W

73. Harel O, Mitchell EM, Perkins NJ, et al. Multiple Imputation for Incomplete Data in Epidemiologic Studies. Am J Epidemiol. 2018;187(3):576–584. doi:10.1093/aje/kwx349

74. Su YS, Gelman A, Hill J, Yajima M. Multiple Imputation with Diagnostics (mi) in R : Opening Windows into the Black Box. J Stat Softw. 2011;45(2). doi:10.18637/jss.v045.i02

75. Lumley T. mitools: Tools for Multiple Imputation of Missing Data. Published online April 26, 2019. Accessed July 22, 2022. https://CRAN.R-project.org/package=mitools

76. VanderWeele TJ, Knol MJ. A Tutorial on Interaction. Epidemiol Methods. 2014;3(1). doi:10.1515/em-2013-0005

77. Bureau UC. 2020 Census Illuminates Racial and Ethnic Composition of the Country. Census.gov. Accessed March 7, 2022. https://www.census.gov/library/stories/2021/08/improved-race-ethnicity-measures-reveal-united-states-population-much-more-multiracial.html

78. McGuire TG, Miranda J. New Evidence Regarding Racial And Ethnic Disparities In Mental Health: Policy Implications. Health Aff (Millwood). 2008;27(2):393–403. doi:10.1377/hlthaff.27.2.393

79. Braveman PA, Cubbin C, Egerter S, et al. Socioeconomic Status in Health Research: One Size Does Not Fit All. JAMA. 2005;294(22):2879. doi:10.1001/jama.294.22.2879

