## Appendix Material for "Racial Differences in Associations Between Adverse Childhood Experiences and Physical, Mental, and Behavioral Health"

**Appendix A.** Key questions used to construct Add Health covariate and outcome measures

| Variable | Add Health Questions | Coding |
|---|---|---|
| Metabolic syndrome | Wave 4 and Biomarker File<br>1. Hypertension<br>o H4SBP: Systolic blood pressure ≥ 130 mmHg, or<br>o H4DBP: Diastolic blood pressure ≥ 80 mmHg, or<br>o H4ID5C: Ever diagnosed with hypertension, or<br>o Current hypertension medication use<br>2. Waist circumference: H4WAIST (> 102 cm for males, > 80 cm for females)<br>3. Triglycerides: TG (top 3 deciles for males, top 2 for females)<br>4. High-density lipoprotein: HDL (bottom 2 deciles for males, bottom 3 for females)<br>5. Diabetes<br>o HBA1C: Glycated hemoglobin (HbA1c) > 5.7%, or<br>o C_MED: Current anti-diabetic medication use (yes/no), or<br>o H4ID5D: Has a doctor, nurse or other health care provider ever told you that you have or had: depression? (yes/no) | 1 = exceeding cutoff in three or more of the five categories<br>0 = exceeding cutoff in fewer than three categories |
| Asthma | H4ID5F (Wave 4): Has a doctor, nurse or other health care provider ever told you that you have or had asthma, chronic bronchitis or emphysema? (yes/no) | 1 = yes<br>0 = no |
| Depression | H3ID26F (Wave 3) Have you taken prescription medication in the past 12 months for depression or stress (yes/no)<br>H4ID5H (Wave 4): Has a doctor, nurse or other health care provider ever told you that you have or had depression? (yes/no) | 1 = yes to either<br>0 = no to both |
| Anxiety | H4ID5J (Wave 4): Has a doctor, nurse or other health care provider ever told you that you have or had: anxiety or panic disorder? (yes/no) | 1 = yes<br>0 = no |
| Suicidal ideation | H4SE1 (Wave 4): During the past 12 months, have you ever seriously thought about committing suicide? (yes/no) | 1 = yes<br>0 = no |

| Variable | Add Health Questions | Coding |
|---|---|---|
| Problematic drug use | H4TO63 (Wave 4): Have you ever taken any prescription drugs that were not prescribed for you, taken prescription drugs in larger amounts than prescribed, more often than pre- scribed, for longer periods than prescribed, or taken prescription drugs that you took only for the feeling or experience they caused? (yes/no)<br>H4TO65C-E (Wave 4): Have you ever used cocaine, crystal meth, or other drugs (such as LSD, PCP, ecstasy, heroin, or mushrooms; or inhalants, not including marijuana or steroids)? (yes/no)<br>H4TO66 (Wave 4): Have you ever injected (shot up with a needle) any illegal drug, such as heroin or cocaine? (yes/no) | 1 = yes to any<br>0 = no to all |
| Parental education | H1RM1 (Wave 1): How far in school did your mother go?<br>H1RF1 (Wave 1): How far in school did your father go?<br>PA12 (Wave 1 Parent): How far did you go in school? | 1 = less than high school, went to trade or vocational school, or GED<br>2 = high school graduate<br>3 = some college<br>4 = college graduate or greater |
| Household size-adjusted income | PA55 (Wave 1): About how much total income, before taxes did your family receive in 1994? Include your own income, the income of everyone else in your household, and income from welfare benefits, dividends, and all other sources.<br>H1HR2 (Wave 1): Please tell me the first names of all the people, other than you yourself, who live in your household. If someone usually lives with you, but is away for a short time, include him or her. | Household income divided by square of household size |

| Variable | Add Health Questions | Coding |
|---|---|---|
| Parental support | H1PF1, H1PF23 (Wave 1): Most of the time, your mother/father is warm and loving toward you (Likert: 1 = not at all, 5 = very much)<br>H1PF4, H1PF24 (Wave 1): You are satisfied with the way your mother/father and you communicate with each other (Likert: 1 = not at all, 5 = very much)<br>H1PF5, H1PF25 (Wave 1): Overall, you are satisfied with your relationship with your mother/father (Likert: 1 = not at all, 5 = very much)<br>H1NM14, H1NF14: How close do you feel to your biological mother/father? (Likert: 1 = not at all, 5 = very much)<br>H1WP10, H1WP14 (Wave 1): How much do you think your mother/father cares about you? (Likert: 1 = not at all, 5 = very much) | Mean of five measures |
| Neighborhood disadvantage score | Wave 1 Contextual File<br>TST90626: Tract proportion of families with income in 1989 below poverty level<br>TST90580: Tract proportion of households with public assistance income<br>TST90680: Tract proportion aged 25+ with no high school diploma or equivalency<br>TST90754: Tract unemployment rate<br>TST90479: Tract proportion of family households that are female householder, no husband present | Mean of five measures |

**Appendix B.** Crosswalk of topical domains, questions in Kaiser ACEs questionnaire, and questions in Add Health used to create ACEs score

| **Domain** | **ACE Questionnaire** | **Add Health Measure Coding** |
|---|---|---|
| Emotional abuse | 1. Did a parent or other adult in the household often...<br>Swear at you, insult you, put you down, or humiliate you?<br>or<br>Act in a way that made you afraid that you might by physically hurt?<br><br>Yes/No | H4MA1 (Wave 4): Before your 18th birthday, how often did a parent or other adult caregiver say things that really hurt your feelings or made you feel like you were not wanted or loved?<br><br>1 = 1-10 times or more<br>0 = this never happened |
| Physical abuse | 2. Did a parent or other adult in the household often...<br>Push, grab, slap, or throw some- thing at you?<br>or<br>Ever hit you so hard that you had marks or were injured?<br><br>Yes/No | H3MA3 (Wave 3): Before the time you started 6th grade, how often had your parents or other adult care- givers slapped, hit, or kicked you?<br><br>1 = 1-10 times or more<br>0 = this never happened |
| Sexual abuse | 3. Did an adult or person at least 5 years older than you ever... Touch or fondle you or have you touch their body in a sexual way?<br>or<br>Try to actually have oral, anal, or vaginal sex with you?<br><br>Yes/No | H3MA4 (Wave 3): Before the time you started 6th grade, how often had your parents or other adult care- givers touched you in a sexual way, forced you to touch him or her in a sexual way, or forced you to have sexual relations?<br><br>1 = 1-10 times or more<br>0 = this never happened |

| Domain | ACE Questionnaire | Add Health Measure Coding |
|---|---|---|
| Emotional neglect | 4. Did you often feel that...<br>No one in your family loved you or thought you were important or special?<br>or<br>Your family didn't look out for each other, feel close to each other, or support each other?<br><br>Yes/No | H1PR8 (Wave 1): How much do you think your family pays attention to you?<br><br>1 = quite a bit - very much<br>0 = not at all - somewhat |
| Physical neglect | 5. Did you often feel that...<br>You didn't have enough to eat, had to wear dirty clothes, and had no one to protect you?<br>or<br>Your parents were too drunk or high to take care of you or take you to the doctor if you needed it?<br><br>Yes/No | By the time you started 6th grade, how often had your parents or other adult care-givers:<br>H3MA1 (Wave 3): Left you home alone when an adult should have been with you?<br>or<br>H3MA2 (Wave 3): Not taken care of your basic needs, such as keeping you clean or providing food or clothing?<br><br>1 = 1-10 times or more for either<br>0 = this never happened |
| Parental separation or divorce | 6. Were your parents ever separated or divorced?<br><br>Yes/No | PA38 - PA54 (Wave 1 Parent)<br>1 = parents divorced or separated before participant turned 18<br>0 = parents not divorced or separated before participant turned 18 |

| Domain | ACE Questionnaire | Add Health Measure Coding |
|---|---|---|
| Mother treated violently | 7. Was your mother or step-mother:<br>Often pushed, grabbed, slapped, or had something thrown at her?<br>or<br>Sometimes or often kicked, bitten, hit with a fist, or hit with something hard?<br>or<br>Ever repeatedly hit over at least a few minutes<br>or<br>Threatened with a gun or knife?<br><br>Yes/No | PB20 (Wave 1 Parent): How much do you fight or argue with your current (spouse/partner)?<br><br>1 = a lot<br>0 = not at all - some |
| Household substance abuse | 8. Did you live with anyone who was a problem drinker or alcoholic or who used street drugs?<br><br>Yes/No | PC49E_2 (Wave 1 Parent): His/her biological father has alcoholism?<br>or<br>PC49E_3 (Wave 1 Parent): His/her biological mother has alcoholism?<br>or<br>H1TO52 (Wave 1): Are illegal drugs easily available to you in your home?<br><br>1 = yes to any<br>0 = no to all |

| **Domain** | **ACE Questionnaire** | **Add Health Measure Coding** |
|---|---|---|
| Mental illness in household | 9. Was a household member depressed or mentally ill or did a household member attempt suicide?<br><br>Yes/No | PA20 (Wave 1 Parent): In general, are you (main parent respondent) happy?<br>or<br>PB16 (Wave 1 Parent): In general do you think (he/she) (main parent respondent's partner or spouse) is happy?<br>or<br>H1SU6 (Wave 1): Have any of your family tried to kill themselves during the past 12 months?<br><br>1 = yes to PA20, yes to PB16, and no to H1SU6<br>0 = any other combination of responses |
| Incarceration of household member | 10. Did a household member go to prison?<br><br>Yes/No | H4WP3 (Wave 4): Has/did your biological mother ever spent/spent time in jail or prison?<br>or<br>H4WP9 (Wave 4): Has/did your biological father ever spent/spend time in jail or prison?<br>or<br>H4WP16 (Wave 4): Has/did your mother figure ever spent/spend time in jail or prison?<br>or<br>H4WP30 (Wave 4): Has/did your father figure ever spent/spend time in jail or prison?<br><br>1 = yes to any<br>0 = no to all |

**Appendix C.** Participant characteristics[a] comparing complete case[b], imputations[c], and missingness[d], stratified by race

| **Characteristic** | **Overall** 12,372 (100%) | **White** 7,742 (73.5%) | **Black** 2,915 (16.9%) | **Asian** 805 (3.18%) | **AI/NA** 76 (0.55%) | **Multiracial** 834 (5.82%) |
|---|---|---|---|---|---|---|
| Male sex | | | | | | |
| Complete case | 5,778 (51%) | 3,672 (51%) | 1,267 (50%) | 418 (53%) | 41 (64%) | 380 (50%) |
| Imputed | 51% | 51% | 50% | 53% | 41% | 50% |
| Missing | 0 (0%) | 0 (0%) | 0 (0%) | 0 (0%) | 0 (0%) | 0 (0%) |
| Age | | | | | | |
| Complete case | 29.0 | 28.9 | 29.2 | 29.2 | 28.8 | 28.8 |
| Imputed | 29.0 | 28.9 | 29.2 | 29.2 | 28.8 | 28.8 |
| Missing | 0 (0%) | 0 (0%) | 0 (0%) | 0 (0%) | 0 (0%) | 0 (0%) |
| Highest parental education | | | | | | |
| Less than high school[e] | | | | | | |
| Complete case | 1,187 (11%) | 685 (9.6%) | 356 (17%) | 69 (13%) | 13 (29%) | 64 (11%) |
| Imputed | 11% | 9.6% | 17% | 13% | 29% | 11% |
| High school diploma | | | | | | |
| Complete case | 3,883 (35%) | 2,520 (34%) | 959 (42%) | 164 (24%) | 19 (34%) | 221 (33%) |
| Imputed | 35% | 34% | 42% | 23% | 34% | 33% |
| Some college | | | | | | |
| Complete case | 2,221 (18%) | 1,405 (19%) | 500 (16%) | 106 (10%) | 19 (24%) | 191 (22%) |
| Imputed | 18% | 19% | 16% | 10% | 24% | 22% |
| College graduate or greater | | | | | | |
| Complete case | 4,897 (36%) | 3,039 (38%) | 1,043 (25%) | 445 (53%) | 24 (14%) | 346 (34%) |
| Imputed | 36% | 38% | 26% | 53% | 14% | 34% |
| Missing | 184 (1.6%) | 93 (1.4%) | 57 (2.5%) | 21 (2.0%) | 1 (<0.1%) | 12 (1.5%) |
| Household income[f] | | | | | | |
| Complete case | 23.3 | 25.2 | 14.2 | 27.3 | 10.6 | 21.2 |
| Imputed | 23.4 | 25.5 | 14.4 | 27.2 | 13.2 | 21.2 |
| Missing | 2,879 (21%) | 1,481 (19%) | 850 (30%) | 338 (40%) | 24 (27%) | 186 (20%) |
| Parental support index (0-5) | | | | | | |

| | | | | | | |
|---|---|---|---|---|---|---|
| Complete case | 3.33 | 3.35 | 3.26 | 3.30 | 3.39 | 3.30 |
| Imputed | 3.33 | 3.35 | 3.26 | 3.30 | 3.38 | 3.30 |
| Missing | 21 (0.2%) | 9 (0.2%) | 9 (0.3%) | 2 (0.3%) | 1 (1.0%) | 0 (0%) |
| Neighborhood disadvantage (0-1) | | | | | | |
| Complete case | 0.14 | 0.12 | 0.23 | 0.13 | 0.23 | 0.15 |
| Imputed | 0.14 | 0.12 | 0.23 | 0.13 | 0.23 | 0.15 |
| Missing | 121 (1.1%) | 80 (1.2%) | 28 (1.1%) | 4 (0.6%) | 0 (0%) | 9 (1.1%) |
| ACEs[g] | | | | | | |
| Elevated (≥ 4) ACEs | | | | | | |
| Complete case | 4,402 (36%) | 3,207 (41%) | 648 (22%) | 260 (32%) | 15 (20%) | 272 (33%) |
| Imputed | 27% | 25% | 33% | 21% | 40% | 35% |
| Summary score (0-10) | | | | | | |
| Complete case | 1.94 | 1.89 | 2.12 | 1.92 | 2.58 | 2.39 |
| Imputed | 2.46 | 2.34 | 2.84 | 2.30 | 3.16 | 2.91 |
| Missing | 7,061 (55%) | 3,899 (50%) | 2,127 (75%) | 499 (59%) | 55 (79%) | 481 (54%) |
| MetS | | | | | | |
| Complete case | 3,131 (29%) | 1,799 (27%) | 852 (38%) | 221 (27%) | 35 (54%) | 224 (31%) |
| Imputed | 29% | 27% | 38% | 27% | 56% | 31% |
| Missing | 1,791 (15%) | 1,009 (14%) | 550 (19%) | 110 (13%) | 12 (17%) | 110 (15%) |
| Hypertension | | | | | | |
| Complete case | 6,082 (51%) | 3,775 (51%) | 1,464 (55%) | 390 (46%) | 46 (68%) | 407 (50%) |
| Imputed | 52% | 51% | 55% | 46% | 66% | 50% |
| Missing | 359 (2.9%) | 192 (2.5%) | 123 (4.2%) | 21 (1.7%) | 5 (12%) | 18 (3.2%) |
| Depression | | | | | | |
| Complete case | 2,195 (19%) | 1,652 (21%) | 296 (10%) | 53 (7.6%) | 11 (14%) | 183 (24%) |
| Imputed | 33% | 35% | 26% | 17% | 32% | 35% |
| Missing | 5,051 (40%) | 2,780 (33%) | 1,507 (54%) | 424 (53%) | 41 (44%) | 299 (35%) |
| Asthma | | | | | | |
| Complete case | 1,867 (15%) | 1,134 (15%) | 453 (15%) | 90 (10%) | 14 (16%) | 176 (24%) |
| Imputed | 15% | 15% | 15% | 10% | 16% | 24% |

| | | | | | | |
|---|---|---|---|---|---|---|
| Missing | 2 (<0.1%) | 2 (<0.1%) | 0 (0%) | 0 (0%) | 0 (0%) | 0 (0%) |
| Anxiety | | | | | | |
| Complete case | 1,518 (13%) | 1,179 (15%) | 182 (6.0%) | 26 (2.9%) | 4 (3.3%) | 127 (18%) |
| Imputed | 13% | 15% | 6.0% | 2.9% | 3.3% | 18% |
| Missing | 2 (<0.1%) | 2 (<0.1%) | 0 (0%) | 0 (0%) | 0 (0%) | 0 (0%) |
| Suicidal ideation | | | | | | |
| Complete case | 845 (7.2%) | 527 (7.1%) | 188 (6.9%) | 48 (6.0%) | 8 (15%) | 74 (10%) |
| Imputed | 7.4% | 7.2% | 7.4% | 6.0% | 15% | 10% |
| Missing | 83 (1.0%) | 29 (0.6%) | 49 (3.2%) | 0 (0%) | 1 (5.7%) | 4 (0.4%) |
| Drug use | | | | | | |
| Complete case | 3,965 (35%) | 3,049 (41%) | 356 (12%) | 197 (25%) | 30 (41%) | 333 (41%) |
| Imputed | 36% | 41% | 12% | 25% | 46% | 42% |
| Missing | 77 (0.8%) | 34 (0.6%) | 35 (1.7%) | 4 (0.4%) | 1 (5.7%) | 3 (< 0.1%) |

Abbreviations: ACE = adverse childhood experience; MetS = metabolic syndrome

[a] Unweighted counts

[b] Weighted proportions (for categorical variables) and means (for continuous) reported

[c] Imputations pooled over 20 datasets; imputation models included all regression variables and variables representing status of parental self-rated health, divorce, employment, disability, retirement, happiness, and welfare receipt, interviewer assessments of neighborhood safety and how well-kept the household is, and number of interruptions to interview with parent

[d] Unweighted counts and weighted proportions

[e] Includes completed vocational school or GED

[f] Equivalence-scaled to adjust for household size

[g] ACE components measured variously across Waves 1, 3, and 4, see Appendix A for details

**Appendix D.** Details and formula for calculation of excess cases

In Chapter 26 of *Modern Epidemiology*, VanderWeele et al[a] introduce the analysis of interaction on both the relative (multiplicative) and absolute (additive) scales using an example where $Y$ is a binary outcome, and $X$ and $G$ are binary predictors. The risk $p$ of the outcome $Y$ when $X = x$ and $G = g$ can be represented by $p_{gx} = P(Y = 1|G = g,\ X = x)$. Risk ratio (RR) and risk difference (RD) effect measures for the "doubly" ($X$ = $G = 1$), and "singly" ($X$ or $G = 1$) exposed can be expressed by:

$$RR_{11} = \frac{p_{11}}{p_{00}}, RD_{11} = p_{11} - p_{00}$$
$$RR_{10} = \frac{p_{10}}{p_{00}}, RD_{10} = p_{10} - p_{00}$$
$$RR_{01} = \frac{p_{01}}{p_{00}}, RD_{01} = p_{01} - p_{00}$$

If we want to estimate additive interaction using measures from a risk ratio regression, we can do so using the interaction contrast ratio or ICR (also called the relative excess risk due to interaction or RERI). The ICR from risk ratio regressions can be expressed as:

$$ICR = RERI_{RR} = \frac{p_{11}}{p_{00}} - \frac{p_{10}}{p_{00}} - \frac{p_{01}}{p_{00}} + 1 = RR_{11} - RR_{10} - RR_{01} + 1$$

If we have estimated risk differences from an additive model, we can calculate a measure of additive interaction by calculating the interaction contrast (IC), which can be expressed and simplified as:

$$IC = RD_{11} - RD_{10} - RD_{01} = p_{11} - p_{10} - p_{01} + p_{00}$$

We can see then that the IC can be recovered from the ICR by multiplying all terms in the ICR by $p_{00}$. The IC is a measure of additive interaction, which can also be scaled up to represent a difference between two groups in number of cases per 100 or 1000 associated with some exposure (see page 37 of VanderWeele and Knol[b]).

[a] VanderWeele TJ, Lash TL, Rothman KJ. Analysis of Interaction In: Modern Epidemiology 4th ed. Lippincott Williams & Wilkins; 2021:619-654.)
[b] VanderWeele TJ, Knol MJ. A Tutorial on Interaction. *Epidemiol Methods*. 2014;3(1). doi:10.1515/em-2013-0005

**Appendix E.** Sensitivity analysis 1: summary ACE score as exposure, Multiracial as referent

**Table E1.** Adjusted[a] within-group RRs and 95% CIs associated with an additional ACE, Add Health 1994-2008

| Outcome | Overall | White | Black | Asian | AI/NA | Multiracial |
|---|---|---|---|---|---|---|
| MetS | 1.00 (1.00, 1.01) | 1.00 (0.99, 1.01) | 1.00 (0.99, 1.01) | 0.99 (0.96, 1.03) | 1.00 (0.94, 1.07) | 1.00 (0.98, 1.02) |
| Hypertension | 1.00 (0.99, 1.00) | 1.00 (0.99, 1.00) | 1.00 (0.99, 1.01) | 1.00 (0.97, 1.03) | 0.96 (0.92, 1.01) | 0.99 (0.97, 1.01) |
| Asthma | **1.01 (1.01, 1.02)** | **1.01 (1.01, 1.02)** | **1.01 (1.00, 1.02)** | 1.00 (0.98, 1.02) | 1.04 (0.99, 1.09) | 1.02 (1.00, 1.04) |
| Depression | **1.04 (1.03, 1.04)** | **1.04 (1.03, 1.04)** | **1.04 (1.02, 1.05)** | 1.03 (1.00, 1.06) | 1.06 (1.00, 1.12) | **1.04 (1.03, 1.06)** |
| Anxiety | **1.01 (1.01, 1.02)** | **1.02 (1.01, 1.02)** | 1.01 (1.00, 1.02) | 1.00 (0.99, 1.01) | 1.00 (0.98, 1.02) | **1.03 (1.02, 1.05)** |
| Suicidal ideation | **1.02 (1.01, 1.02)** | **1.01 (1.01, 1.02)** | 1.01 (1.00, 1.02) | **1.03 (1.01, 1.05)** | 1.02 (0.97, 1.08) | 1.02 (1.00, 1.03) |
| PDU | **1.03 (1.02, 1.04)** | **1.03 (1.03, 1.04)** | **1.02 (1.01, 1.03)** | 1.03 (1.00, 1.06) | **1.08 (1.04, 1.12)** | **1.04 (1.03, 1.06)** |

**Table E2.** Exponentiated interaction[b] term betas and 95% CIs from regression models, Add Health 1994-2008

| Outcome | White | Black | Asian | AI/NA | Multiracial | p-value |
|---|---|---|---|---|---|---|
| MetS | 1.00 (0.98, 1.02) | 1.00 (0.98, 1.02) | 1.00 (0.96, 1.04) | 1.00 (0.94, 1.07) | (ref.) | 0.99 |
| Hypertension | 1.01 (0.99, 1.02) | 1.01 (0.99, 1.03) | 1.01 (0.98, 1.05) | 0.97 (0.93, 1.02) | (ref.) | 0.46 |
| Asthma | 0.99 (0.97, 1.01) | 0.99 (0.97, 1.01) | 0.98 (0.95, 1.01) | 1.02 (0.97, 1.08) | (ref.) | 0.49 |
| Depression | 0.99 (0.98, 1.01) | 0.99 (0.98, 1.01) | 0.99 (0.95, 1.02) | 1.01 (0.95, 1.08) | (ref.) | 0.90 |
| Anxiety | 0.98 (0.97, 1.00) | **0.98 (0.96, 0.99)** | **0.97 (0.95, 0.98)** | **0.97 (0.95, 0.99)** | (ref.) | ***** < 0.001** |
| Suicidal ideation | 1.00 (0.98, 1.01) | 0.99 (0.98, 1.01) | 1.01 (0.98, 1.03) | 1.00 (0.95, 1.06) | (ref.) | 0.82 |
| Drug use | 0.99 (0.98, 1.00) | **0.98 (0.97, 0.99)** | 0.99 (0.95, 1.02) | 1.04 (1.00, 1.08) | (ref.) | *** 0.01** |

**Table E3.** Excess[b] cases[c] per 1,000 and 95% CIs[d] associated with an additional ACE, Add Health 1994-2008

| Outcome | White | Black | Asian | AI/NA | Multiracial |
|---|---|---|---|---|---|
| MetS | 2 (-17, 21) | 3 (-18, 24) | -3 (-41, 32) | 4 (-74, 58) | (ref.) |
| Hypertension | 9 (-10, 30) | 14 (-7, 36) | 13 (-26, 50) | -36 (-108, 29) | (ref.) |
| Asthma | -14 (-40, 13) | -13 (-42, 15) | -30 (-68, 7) | 27 (-35, 86) | (ref.) |
| Depression | -5 (-20, 11) | -9 (-26, 8) | -17 (-49, 13) | 11 (-48, 61) | (ref.) |
| Anxiety | -17 (-33, 2) | **-25 (-41, -7)** | **-37 (-55, -19)** | **-35 (-58, -13)** | (ref.) |
| Suicidal ideation | -4 (-20, 12) | -6 (-25, 13) | 6 (-20, 31) | 6 (-55, 56) | (ref.) |
| Drug use | -19 (-44, 7) | **-45 (-74, -17)** | -29 (-96, 30) | 62 (4, 113) | (ref.) |

**Commented [RCA1]:** Methods mention running, crude, adjusted and stratified models but the results only show the latter, so update the methods description to remove discussion of other models if don't consider or present them.

**Commented [RCA2]:** I am having trouble squaring how this number is > 1 (indicating that the model is super-multiplicative) while in the next table the RERI in the same cell is <0, indicating that the model is sub-additive. That is impossible…. Is there something I am missing??

I think I am also struggling with Table 1.3 being race stratified (without whites as a referent group) but Table 1.4 having whites as the referent group. Maybe this is partially explaining this counter intuitive finding. Is it possible to make a table that would go in a supplement that is for relative interaction with Whites as the referent group – and then can see if the interactions agree (eg if something is super multiplicative than it is super additive, and if it sub additive then it is also sub multiplicative).

Perhaps a question for Patrick.

**Commented [RCA3]:** Can these be scaled to make them more interpretable? For example, is this like 13 more cases of htn per 1000 Black participants vs. white participants? Would recommend multiplying whole table by 1000 and indicating interpretation as excess cases per 1000.

**Commented [TLH4R3]:** Great suggestion. These are so much more interpretable now. It also makes me realize how wildly huge some of these estimates/CIs are

Abbreviations: CI = confidence interval; ACE = adverse childhood experience; MetS = metabolic syndrome
Boldface indicates statistical significance (95% CI does not include the null, or global Wald test for interaction * $p < 0.05$, ** $p < 0.01$, ***$p < 0.001$)
[a] Models adjusted for participant age, sex, parental education, household size-adjusted income, parental support, and neighborhood disadvantage score
[b] Race × summary ACEs score interaction, Multiracial as referent group
[c] 95% confidence intervals calculated from 10,000 resamples of interaction contrast

**Appendix F.** Sensitivity analysis 2: elevated ACEs as exposure, White as referent

**Table F1.** Exponentiated interaction[a] term betas and 95% CIs from regression analyses, Add Health 1994-2008

| Outcome | White | Black | Asian | AI/NA | Multiracial | p-value |
|---|---|---|---|---|---|---|
| MetS | (ref.) | 1.01 (0.95, 1.06) | 1.00 (0.87, 1.14) | 1.00 (0.79, 1.28) | 1.03 (0.94, 1.12) | 0.98 |
| Hypertension | (ref.) | 1.02 (0.97, 1.07) | 1.03 (0.91, 1.17) | 0.87 (0.70, 1.08) | 0.99 (0.92, 1.06) | 0.65 |
| Asthma | (ref.) | 0.99 (0.94, 1.04) | 0.97 (0.89, 1.05) | 1.11 (0.90, 1.37) | 1.08 (1.00, 1.18) | 0.21 |
| Depression | (ref.) | 1.00 (0.95, 1.05) | 0.98 (0.86, 1.10) | 1.07 (0.80, 1.42) | 1.02 (0.94, 1.09) | 0.97 |
| Anxiety | (ref.) | 0.98 (0.94, 1.02) | **0.93 (0.89, 0.96)** | 0.94 (0.87, 1.02) | 1.06 (0.98, 1.14) | ***** < 0.001** |
| Suicidal ideation | (ref.) | 1.00 (0.95, 1.04) | 1.03 (0.95, 1.12) | 1.02 (0.79, 1.31) | 1.00 (0.93, 1.07) | 0.96 |
| Drug use | (ref.) | 0.97 (0.93, 1.02) | 0.98 (0.86, 1.12) | 1.18 (0.95, 1.45) | 1.02 (0.96, 1.09) | 0.29 |

**Table F2.** Excess[a] cases[b] per 1,000 and 95% CIs[c] associated with elevated ACEs, Add Health 1994-2008

| Outcome | White | Black | Asian | AI/NA | Multiracial |
|---|---|---|---|---|---|
| MetS | (ref.) | 6 (-45, 56) | -1 (-114, 117) | 3 (-238, 268) | 26 (-50, 109) |
| Hypertension | (ref.) | 18 (-39, 74) | 35 (-99, 175) | -163 (-394, 97) | -14 (-96, 62) |
| Asthma | (ref.) | -18 (-90, 51) | -46 (-148, 60) | 147 (-138, 491) | **123 (8, 252)** |
| Depression | (ref.) | -13 (-75, 49) | -47 (-179, 96) | 80 (-251, 511) | 17 (-74, 112) |
| Anxiety | (ref.) | -29 (-74, 15) | **-91 (-140, -47)** | -72 (-171, 17) | 71 (-27, 164) |
| Suicidal ideation | (ref.) | -2 (-50, 46) | 34 (-58, 128) | 27 (-265, 346) | -3 (-88, 83) |
| Drug use | (ref.) | **-97 (-194, -2)** | -72 (-337, 225) | 409 (-98, 925) | 50 (-93, 200) |

Abbreviations: CI = confidence interval; ACE = adverse childhood experience; MetS = metabolic syndrome

Boldface indicates statistical significance (95% CI does not include the null, or global Wald test for interaction * $p < 0.05$, ** $p < 0.01$, ***$p < 0.001$)

[a] Race × elevated ACEs score interaction, White as referent group

[b] Rounded to the nearest whole person

[c] 95% confidence intervals calculated from 10,000 resamples of interaction contrast
